# RADAR-IoT: An open-source, interoperable and extensible IoT gateway framework for healthcare and beyond

**DOI:** 10.1101/2023.08.25.23294420

**Authors:** Yatharth Ranjan, Jiangeng Chang, Heet Sankesara, Pauline Conde, Zulqarnain Rashid, Shaoxiong Sun, Richard Dobson, Amos Folarin

**Affiliations:** King’s College London; National University of Singapore; King’s college London; Kings College London

**Keywords:** IoT, Gateway, Healthcare, Radar-Base, Extensible

## Abstract

IoT sensors offer a wide range of sensing capabilities, many of which have the potential for health or medical applications. Existing solutions for IoT in healthcare have notable limitations, such as limited I/O protocols, limited cloud platform support, and limited extensibility. Therefore, the development of an open-source Internet of Medical Things (IoMT) gateway solution that addresses these limitations and provides reliability, broad applicability, and utility would be highly desirable. Combining a wide range of sensor data streams from IoT devices with ambulatory mHealth data would open the potential for providing a detailed 360-degree view of the relationship between patient physiology, behaviour, and environment. To harness this potential, RADAR-IoT has been developed as an open-source IoT gateway framework. Its purpose is to connect multiple IoT devices at the edge, perform on-device data analysis, and integrate with cloud-based mobile health platforms such as RADAR-base, enabling real-time data processing.

## I. Introduction

### A. Internet of Things

The current landscape is witnessing a significant expansion in modern sensing capabilities and network-connected devices. This growth presents an opportunity to collect largescale health data continuously and remotely, facilitating timely analysis for the identification and monitoring of individuals’ health status. Within the market, there exist numerous established and emerging mobile health (mHealth) and Internet of Things (IoT) platforms. These platforms have gained increasing importance due to technological advancements and the widespread integration of wearables into daily life. As a result, unprecedented possibilities (e.g. home-based and ambulatory monitoring and just-in-time interventions) have emerged in both research and clinical applications.

The Internet of Things can be understood as the network of physical objects—“things”—that are comprised of sensors, software, and other technologies for the purpose of interacting data with other devices and systems over the Global Internet. The Internet of Things represents a wide category of devices and sensors with the common attribute of network connectivity, they form a superset that includes wearable devices and smartphones. An IoT gateway device acts as a hub and interconnects all IoT devices at a location (called at-edge, i.e. near the source of the data) to the cloud-based platforms and IoT devices in other locations. Its functionality may also include rudimentary processing of the data and security- and privacy-oriented access. With recent developments such as neural processing units (NPUs) for advanced AI and 64-bit CPUs at the edge, this scenario becomes increasingly realistic and exciting.

Surveys on the state of IoT in healthcare [1], [2] reveal that most IoT platforms suffer from limitations such as limited sensor support, closed-source systems, limited I/O protocols, no proper cloud platform support, no demonstrable proof of concept, and limited extensibility. To address these issues, we designed an open-source Internet of Things (IoT) gateway framework that can interconnect multiple IoT devices on the edge, perform rudimentary analysis, and integrate well with established mHealth platforms in the cloud. Finally, we evaluated these capabilities in a proof-of-concept deployment.

### B. Motivation and Related Work

The Internet of Medical Things (IoMT) sector of the healthcare market is projected to reach USD 534.3 billion by 2025, expanding at a compound annual growth rate (CAGR) of 19.9% [3] as shown in Figure 1. In 2018, the market was valued at USD 147.1 billion. This significant growth can be attributed to the proliferation of wearables and IoT-enabled devices into daily life, such as smart virtual assistants and smart lighting, as well as the emergence of cloud computing, which enables the management and deployment of millions of connected IoT devices. However, uptake of these technologies in the medical/healthcare domain is more attenuated with more than 50% of doctors expressing concerns over IoT devices reliability [4]. This is due to the fact that present IoT data is often unprocessed, non-standardized, and lacks actionable or valuable intelligence. The integration of data from multiple devices with a mHealth platform capable of analysing realtime data at both the network edge and in the cloud holds the potential for the generation of actionable information, with potentially large variety of sensors enabling more use-cases and scenarios. Leveraging this combined data processing capability, combined with appropriate validation, significant advancements can be achieved in the realm of healthcare with a detailed 360-degree view of the patient’s physiology, behavior, and environment.

**Fig. 1:**
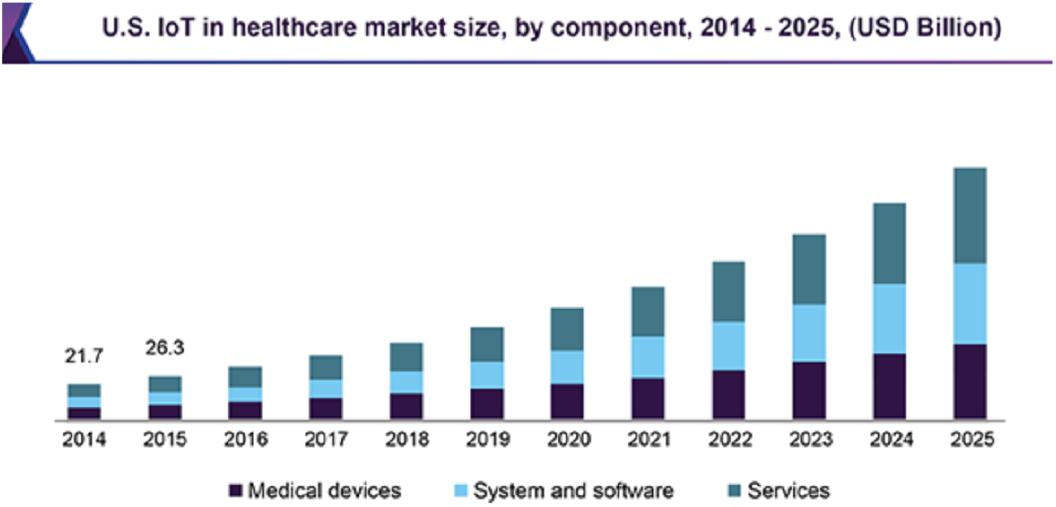
The market trend of the Internet of Medical Things (IoMT) [3]

While there is a wide range of IoT platforms available in the market, most of them are commercial or closed-source, and even open-sourced platforms are typically consumer-oriented, such as those designed for controlling smart home devices. There is however a shortage of open-source mHealth platforms that allow for interconnected IoT devices and offer a common interface for collecting, aggregating, and analyzing real-time data from multiple sources. These sources include wearables, IoT sensors, mobile devices, and medical devices, which are essential for research and clinical domains in healthcare.

Various IoT gateway frameworks are compared in Table I. RADAR-IoT [16] takes inspiration from such frameworks for its design and does not mean to be an alternative or replace them, rather it tries to extend them, by either having the potential to connect to them to collect the data, or, to pass data to them, hence making use of existing functionalities of these platforms. While having generalized platforms like these are good for broader use cases, these do not shine for very specific use cases, for instance, healthcare and remote monitoring of patients. Most of the platforms typically use an internal message bus, like D-Bus, but the benefits of having an external industry-standard message broker are many-fold, e.g. event-driven programming and analysis, language and platform-agnostic publishers and subscribers, enormous extensibility and interoperability. Some platforms are primarily cloud-based (these can only function with a connection to their respective cloud servers), this is a significant limitation where a local network is preferred in privacy-constrained environments such as hospitals. A Smart Gateway Framework for IoT Services [7] and A Novel Cognitive IoT Gateway Framework [9] focus on high-level context abstraction, cognition and knowledge. While our target framework does not focus on this in the first instance, it has high levels of abstraction to enable incorporation of such contexts, cognition and ontologies into the framework in the future. The primary objective of the proposed IoT framework was to enhance the existing RADAR-Base mHealth platform by incorporating IoT sensor capabilities, while also ensuring the framework’s extensibility to accommodate future use cases and concepts. This integration would unlock the potential to combine diverse sensor data streams from IoT devices with ambulatory mHealth data and gain deeper insights into the complex interplay between patients’ physiology, their behavior and well-being, and the surrounding environment. This can lead to more informed healthcare decisions and personalized interventions.

**TABLE I:**
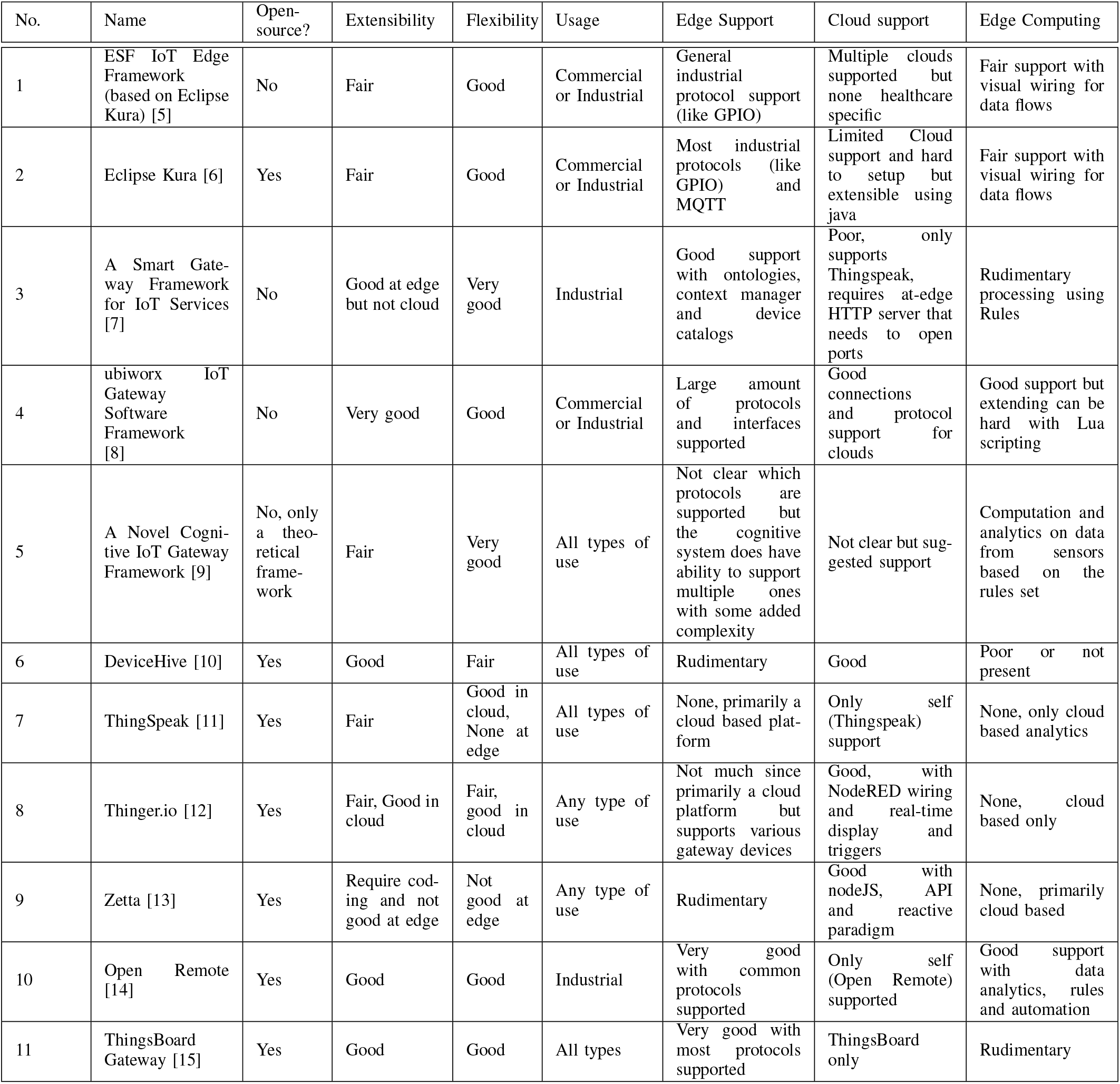
IoT gateway frameworks comparison.

The IoT environment is ubiquitous, making it impractical to impose standards and expect universal compliance. The large-scale network and heterogeneity of things in IoT domain, along with the large number of events that can be generated by these IoT devices, pose new challenges in application development. These challenges make the development of solutions in ubiquitous computing considerably more difficult [17]. Middleware can offer common services for applications, integrate heterogeneous computing and communication devices, and support interoperability within diverse applications and services running on these devices. Essentially functioning as hidden translation layer, middleware enables communication and data management for distributed applications. RADAR-IoT aims to be a middleware to overcome such challenges.

Existing IoT gateways shown in Table I are mostly commercial or industrial. Some healthcare and research organizations provide gateways as physical devices [18], limiting their adoption and control. Kesavan et al [19] proposed a gateway framework for smart health, but it is limited to a few body-attached sensors, supports only Bluetooth and Wi-Fi, and limited info is provided on cloud platform integration.

### C. RADAR-Base

RADAR-base (Remote Assessment of Disease And Relapses) [20] is an open-source platform that integrates data streams from various wearables and mobile technologies to collect sensor data in real-time and store, manage and share the collected data, knowledge and insights with researchers for analysis and actionable intelligence. As shown in Figure 2, it supports both passive and active data collection through two applications, pRMT and aRMT, which monitor movement, location, audio, calls, texts, and app usage, and include questionnaires to gather patient information. RADAR-base has been and is currently being used in various research studies [21], [22], [23], [24], [25], [26], [27], [28], [29], [30], [31], [32], focusing on personal sensing. However, the platform’s integration with wearable devices is limited by vendor availability of SDKs and REST APIs. RADAR-IoT extends the platform to include IoT sensors enabling support of a wider range of use cases.

**Fig. 2:**
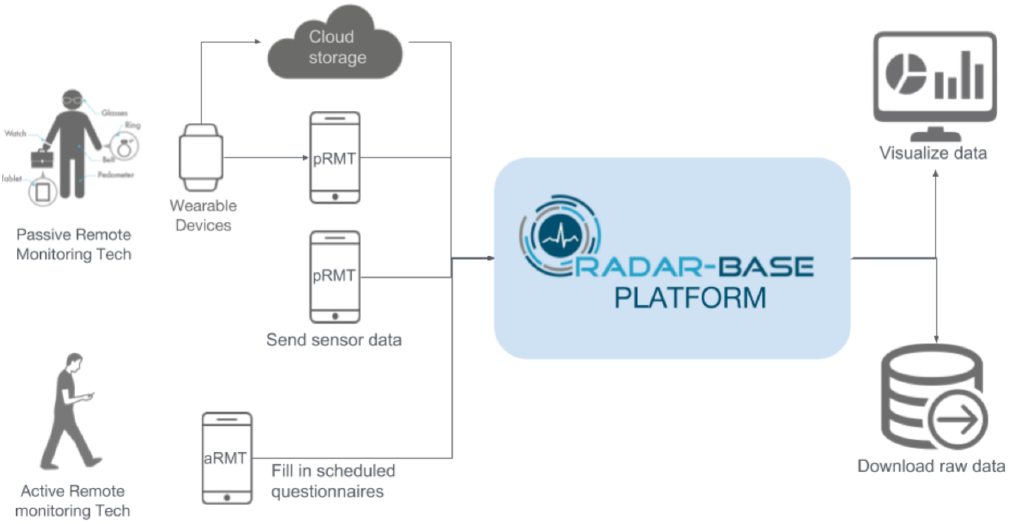
The overview of Radar-base [20]

### D. Design Requirements

Several challenges were considered here due to the diverse nature of the IoT ecosystem as they influence the needs and design decisions for the IoT Gateway module as mentioned in the survey [17]. An IoT edge framework is proposed with design informed based on the following:

1. Extensibility and flexibility: allow integration of any sensor or source and destinations.
2. Modularity: allow for services/components to communicate through a common API, allowing them to be augmented or replaced by custom implementations facilitating re-usability.
3. Support a Publish/Subscribe design pattern [33]: allows local-level interoperability and scalability at the edge.
4. Allow data can be consumed, processed and uploaded in several ways flexible enough to accommodate new use cases and requirements.
5. Operate on low-power and resource-edge devices (like raspberry pi zero [34]).
6. Reliability: the system should allow continuous monitoring from a host of different sensors with different hardware interfaces with data standardisation.
7. Data analysis: including AI applications and machine learning at the edge.
8. Enable remote deployment and orchestration of services at scale.
9. Provide for interoperability with other IoT devices complying with a subset of the OSGI standard [35].
10. Support service and machine discovery at the edge.
11. Designed using modern technologies to get the best-in-class features and performance.

## II. Architecture

RADAR-IoT (RADAR-base - Internet of Things) is a flexible and extensible Internet of Things edge gateway [36] framework for connecting a wide variety of sensors and provision of flexible data processing of those inputs and support for uploading IoT sensor data to RADAR-base platform. RADAR-IoT also supports uploading to InfluxDb [37] (local or cloud) and provides real-time visualization on Grafana [38] dashboards and can be easily extended to include more cloud destinations. All the components are based on abstract base components, streamlining the addition of more data processing variants and sensors. The use of an external industry standard publish/subscribe [33] platform like Redis [39] or MQTT [40] makes the RADAR-IoT framework language and platform agnostic.

### A. Architecture Overview

The architecture of the RADAR-IoT framework, as shown in Figure 3, illustrates its components and data flows from left to right, culminating in the transmission of data to external services. The framework is very flexible and compatible with various IoT sensors (e.g. environment sensors, video and audio) through standard Input/Output protocols (like I2C, GPIO, Serial and more). Data points are validated using a schema and converted into a format suitable for downstream components. These messages are then forwarded or published to the publisher-subscriber platform, where multiple consumers can access them. One such consumer reads these messages and uploads them to external services such as the dashboard and the RADAR-Base platform [20]. The detailed architecture of the framework, depicted in Figure 4, portrays its various layers and components. Once data is published, it can be used in diverse ways, including cloud transmission, on-device data analysis or processing, and Node-Red [41] flows (which is further discussed below).

**Fig. 3:**
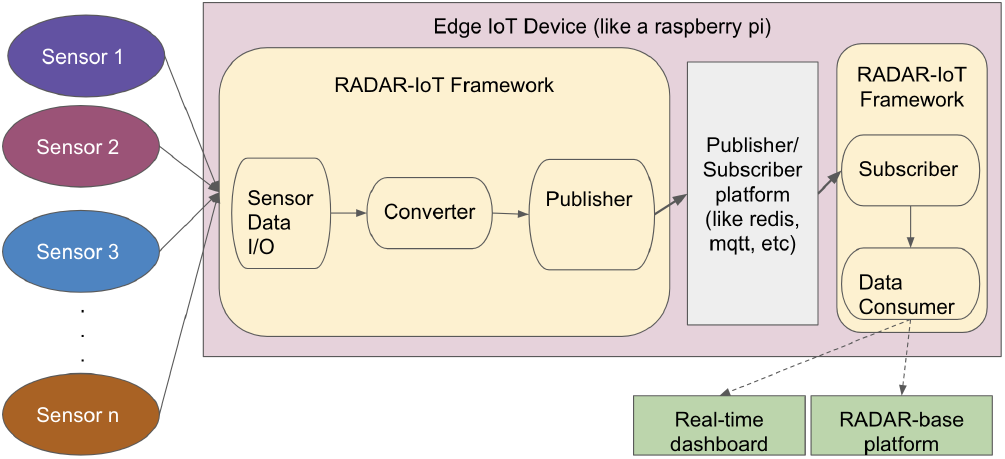
Architecture Overview of RADAR-IoT

**Fig. 4:**
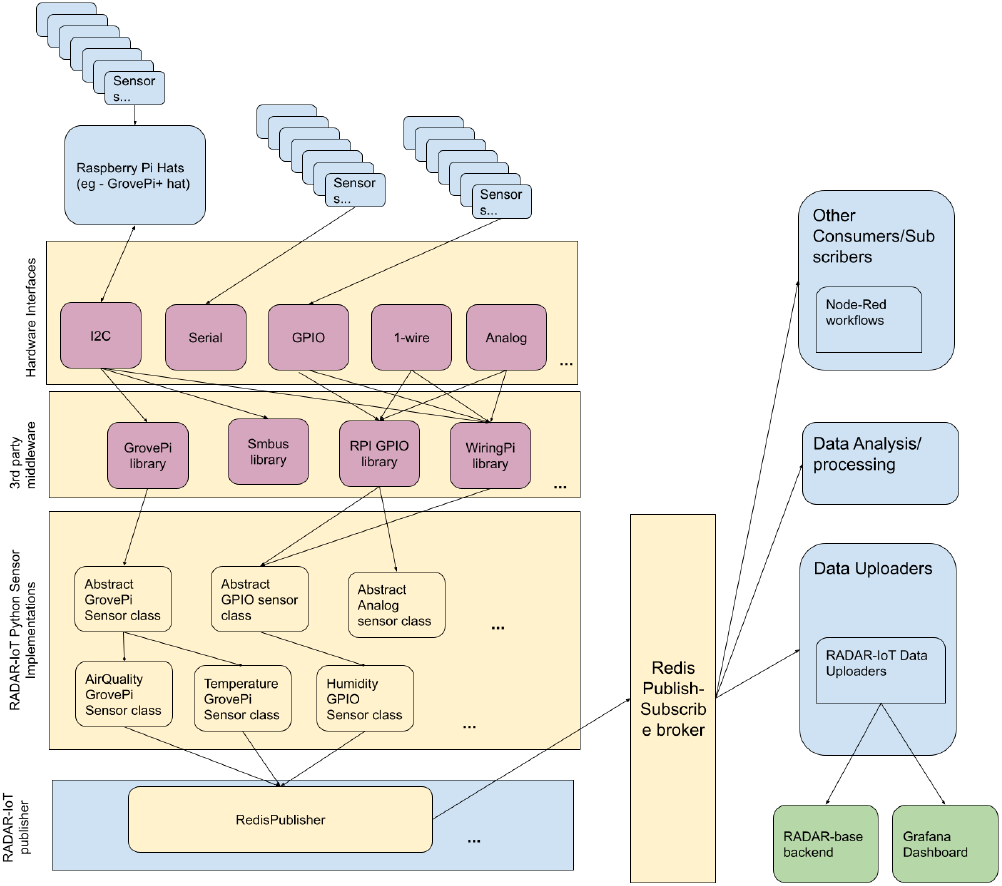
Detailed architecture of RADAR-IoT

### B. Publish/Subscribe Broker or Message Queue

RADAR-IoT uses a Publish-Subscribe (pub-sub) broker [33] as a message queue for publishing and consuming sensor data, providing several benefits. Firstly, an external and widely used pub-sub system (such as Redis [39], Kafka [42], or MQTT [40]) enhances reliability over creating a custom internal message bus, allowing direct data syncing to the cloud running the same pub-sub broker. Secondly, the framework enables bi-directional production and consumption of data between the data-consumer and external applications or services. This also allows for multiple types of publishers, such as broadcasting data from one device to another anywhere on the same network. Thirdly, event-driven programming and analysis are supported. Lastly, publishers and subscribers are language and platform-agnostic.

### C. OGC Sensor Things API

The Open Geospatial Consortium OGC SensorThings API provides an open, geospatial-enabled and unified way to interconnect the Internet of Things (IoT) devices, data, and applications over the Web [35]. A subset of specifications from OGC API was used to design the RADAR-IoT framework.

The channel or topic names in the Publish/Subscribe system provide a standard to specify the task, hence providing a standardised way to publish and consume data. Using this approach canonically named endpoints for controlling and configuring the sensors can be exposed. For instance, for a sensor with *<*sensor-name *>*we use the following format for various endpoints

- /sensor/*<*sensor-name*>*/data-stream
- /sensor/*<*sensor-name*>*/error
- /sensor/*<*sensor-name*>*/control
- /sensor/*<*sensor-name*>*/config

We have exposed an endpoint that contains comprehensive metadata from the sensors in operation, essentially making the system self-describing.

### D. State Machine

A state machine, also known as a finite state machine, is a mathematical model used to describe the behavior of systems. It consists of a set of states, transitions between those states, and actions. In a state machine, the system is always in one of a finite number of states, and the transitions between those states are triggered by events or conditions. Incorporating a state machine [43] into the sensor abstractions for the purpose of capturing and monitoring sensor lifecycle events confers enhanced visibility and insight into the framework and sensors, thereby enabling more resilient fault isolation, diagnosis, and efficient resolution. The resulting events and logs of a particular state are also disseminated to the pub-sub system, thereby enabling downstream and upstream consumers and applications to leverage them for a diverse range of tasks.

### E. Abstraction

The RADAR-IoT framework leverages interfaces and abstract classes to a significant extent. The adoption of object-oriented programming principles [44] grants the system enhanced flexibility and extensibility. Among the specific advantages are the following:

- The majority of the components are abstracted and reusable. The Sensor abstract class serves as the fundamental unit for collecting sensor data, with all sensor implementations required to extend this class or one of its subclasses.
- The framework facilitates the creation of new abstractions based on existing ones.
- The system’s extensibility, encapsulation and reusability are augmented.
- Reliable incremental testing is enabled, as only the specific extension needs to be unit tested, given that the abstractions have already been tested.

### F. Security and Privacy

When collecting health and environmental data, security and privacy are paramount concerns, particularly given the substantial number of endpoints in IoT systems. To address these issues, RADAR-IoT incorporates several measures. The gathering of sensor data happens over a secure channel accessible only through hardware interfaces. After data is gathered from the sensor, it is transmitted to the publish/subscribe broker, which can be configured with a strong username and password to ensure that publishers and subscribers authenticate before accessing or submitting data. Additionally, all data is associated with pseudonymised identifiers to safeguard privacy and prevent user or environment identification. In cases where identifiable data such as audio is involved, on-device feature extraction is performed to extract non-reversible features which can be published in place of the raw audio. This feature extraction capability is integrated into the RADAR-IoT framework as a module. Finally, when uploading data to the cloud, RADAR-IoT employs the standard security mechanisms of the target systems. For instance, the RADAR-Base system employs the OAuth 2.0 [45] industry standard for securing data at REST.

### G. Data Typing and Schematisation

Data typing, validation and standardization are provided with the help of **Avro Schemas** [46]. The data collected from the sensors is validated against a given Avro schema based on sensor type and only then it is converted to be published, this helps standardise the data and manage schema evolution and backward compatibility. The converter can be configured to read schemas from a variety of different sources including the local filesystem, hosted files on GitHub and from the Confluent Schema Registry component [47]. The converter can also be configured to disable validation if it is not required. The data accepted by the RADAR-Base mHealth platform also needs to be in Avro format and hence the final step also includes schema validation for the data. If the data is not compatible with the schema, the data will not be accepted for publishing.

### H. Methods of Deployment

The RADAR-IoT platform is modular and comprises 3 different components that need to be deployed:

1. The Python module connects to Sensors and handles I/O. It also validates the data using Avro Schemas [46] and converts it into a more useful format before publishing to the pub-sub broker.
2. The Publish-Subscribe Broker (current options include Redis [39], MQTT [40])
3. The data consumers, this module can be used to upload data to the cloud, on-device analytics and more.

By packaging the framework as Docker images we can use orchestration frameworks like Nebula [48] for deploying, maintaining and upgrading (remotely) on a very large number of IoT devices with consistency and reliability.

### I. Methods for Management, Maintenance and Monitoring

The management and orchestration of services in an IoT platform an important aspect of non-trivial deployments, especially if the platform needs to scale to hundreds of thousands of devices. A common scenario is having a large number of devices and needing to update the version of the sensor interface service on all of them, doing this manually on each of those (even remotely) is not feasible. RADAR-IoT addresses this through automation using external open-source solutions for management, maintenance and monitoring. These include:

- Management and maintenance: Nebula [48] (an orchestration tool for IoT devices that use Docker-based services) and Dataplicity [49] (a remote IoT device management service)
- Monitoring: Node-Red [41] + Grafana [38] (providing inter- and intra-device monitoring and alerting) and Netdata [50] (professional system monitoring)

## III. Deployment Configurations

The RADAR-IoT framework’s modular architecture allows for deployment in various configurations to meet diverse use cases. Some typical patterns are discussed below.

### A. RADAR-IoT Configurations

#### 1) Multiple Master Devices

This configuration, as depicted in Figure 5(a), allows for one or multiple autonomous IoT gateway devices running RADAR-IoT to gather data from sensor sets. These devices operate independently and do not communicate with each other.

**Fig. 5:**
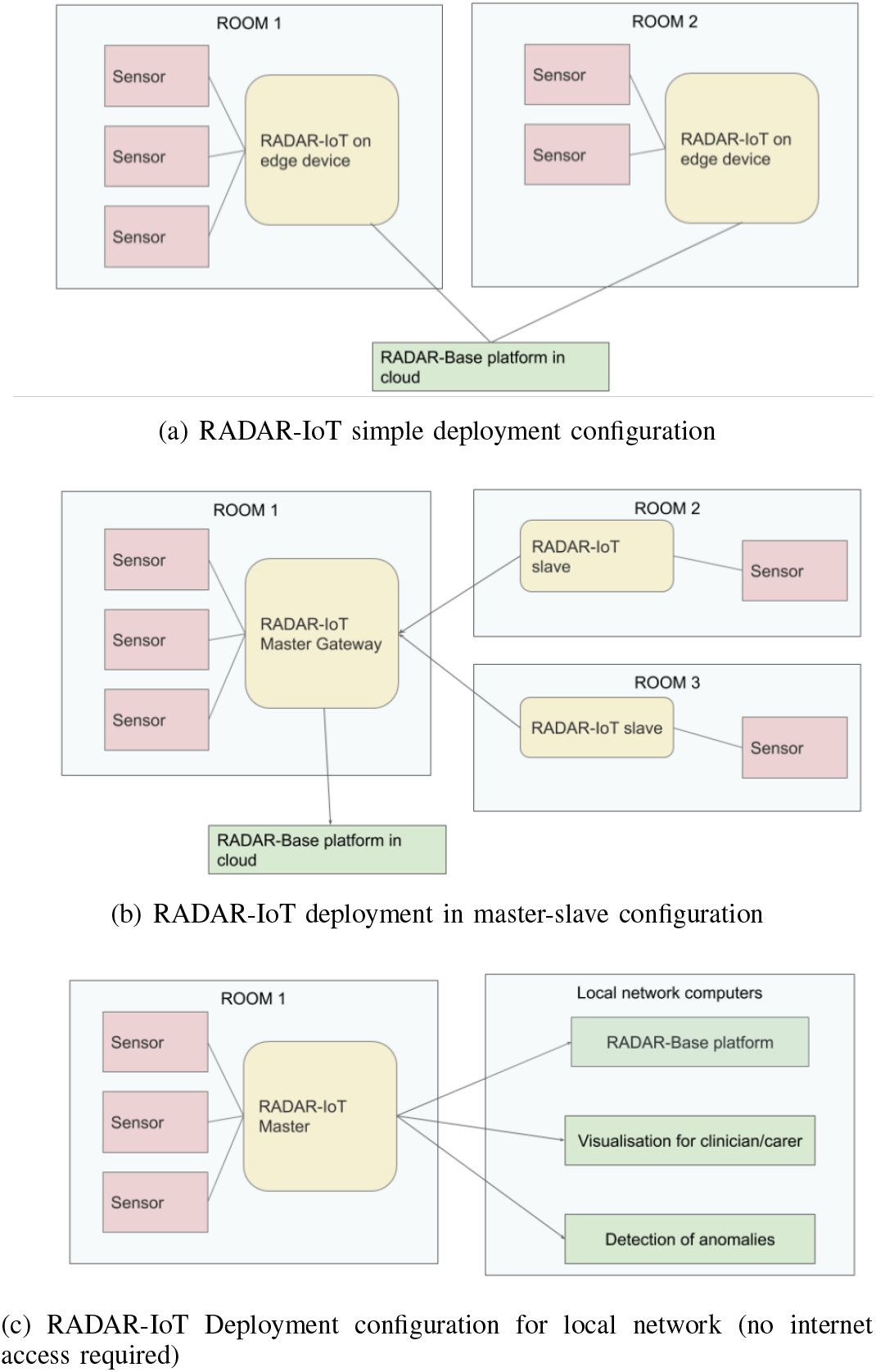
Deployment samples

#### 2) Master and Slaves

The proposed configuration, as illustrated in Figure 5(b), involves the utilization of a master-slave architecture within the RADAR-IoT framework. In this setup, a single master device acts as the primary gateway and hosts all components of the framework. Multiple slave devices equipped with sensors are responsible for interfacing with and collecting data from the sensors. These slave devices transmit the collected data to the master device. The master device can also directly interface with and collect data from the sensors if needed. However, only the master device interacts with data uploading and consuming services, such as dashboards and cloud platforms.

This configuration offers several advantages, especially in situations where deploying multiple independent IoT gateway devices is not feasible or resource constraints exist. By employing a master-slave architecture, data collection and management are optimized while minimizing resource utilization. This approach ensures efficient and centralized data processing while accommodating limitations in terms of available resources or deployment feasibility.

#### 3) Local Network Only

While deployment configurations a) and b) typically utilise a remote or cloud-hosted RADAR-base platform, it is also possible to deploy RADAR-base on the local network servers and so satisfy criteria for local deployment. This configuration precludes internet access, which is typically designated for in-hospitals where transmitting data outside the internal network is not desired. Since all RADAR-IoT components and associated external services are open-source, they can be deployed on local network computers/servers and operate effectively in a completely isolated environment. The example configuration depicted in Figure 5(c) comprises the RADAR-Base platform, the Grafana dashboard for visualization, and the AI/ML data consumer for anomaly detection, all of which are supported by RADAR-IoT.

### B. Node Red Workflows

Node-RED is a low-code programming tool that enables the integration of hardware devices, APIs, and online services in novel and innovative ways. It features a browser-based editor that facilitates the creation of flows using a diverse range of nodes from the palette, which can be deployed to its runtime with a single click [41]. Node-RED is capable of generating a large number of flow combinations for various use cases, as demonstrated by an example use case of alerting in Figure 6. To read data from RADAR-IoT, only the Redis-In or MQTT node (based on the pub-sub broker in use) is required. The example in 6 illustrates an alert workflow that triggers an email if no data is received within a specified time window, such as two hours. This feature enables administrators to investigate potential issues with data collection. Another flow demonstrated is reading audio from MQTT broker, performing feature extraction and then uploading it to the cloud.

**Fig. 6:**
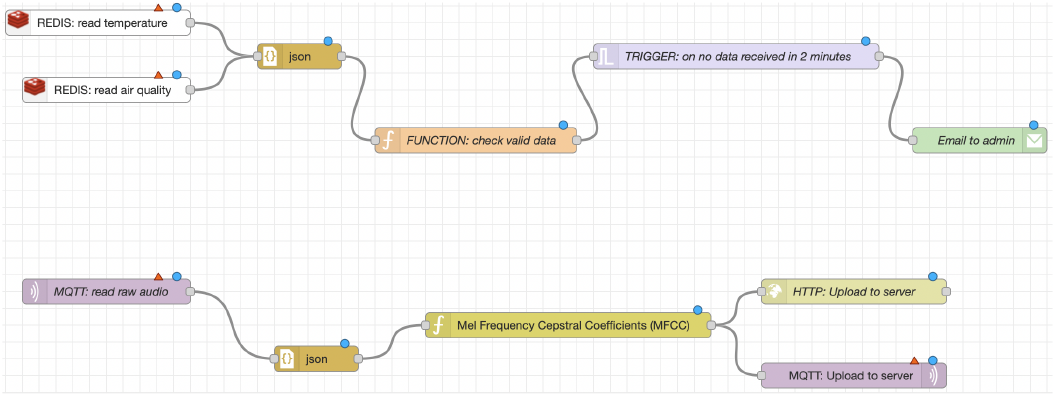
Example Node-Red workflow augmenting RADAR-IoT

This potentially makes the framework extremely extensible to support a very wide variety of outputs with the help of nodes and flows.

### C. Connection to Other Cloud IOT Platform Backends

The RADAR-IoT framework is highly flexible and supports the Grafana dashboard and RADAR-Base platform. Moreover, it can be easily extended to multiple cloud backends, as demonstrated in Figure 7. For instance, third-party IoT cloud platforms such as Google Cloud IoT [51] can be seamlessly integrated with the RADAR-IoT gateway framework due to the publish-subscribe broker feature that most IoT platforms offer.

**Fig. 7:**
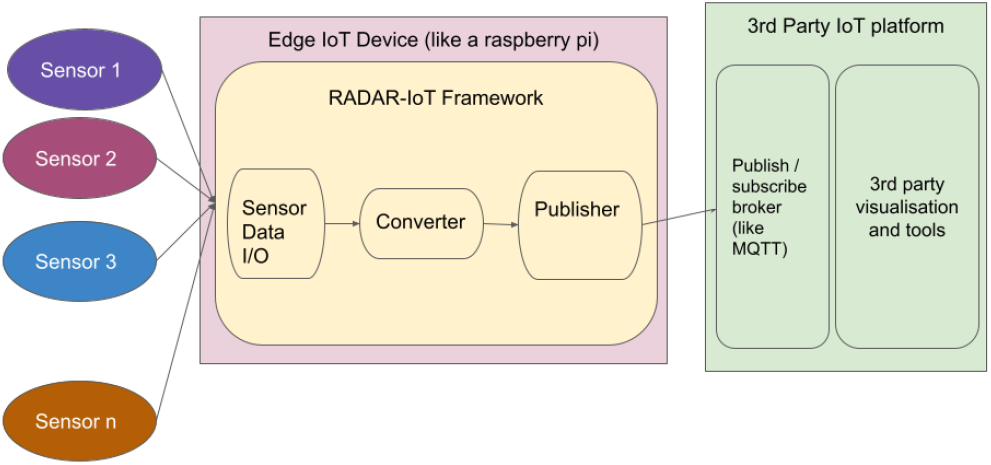
RADAR-IoT use-case with a third-party cloud IoT platform like Google Cloud IoT [51]

## IV. PoC Demonstration

### A. Aims

A Proof-of-Concept (PoC) demonstration of the RADAR-IoT framework is discussed here to illustrate the following objectives:

- Evaluate and demonstrate the framework in a real-world setting.
- Establish that the integration framework and the sensors perform as expected.
- Collect some real-world sensor data with positive control test conditions to assess changes in state to a known perturbation.

### B. Methods

For this study, the RADAR-IoT framework was deployed in two distinct settings: a shared office space and a home-based environment, using a Raspberry Pi 4B device [34] equipped with four types of sensors - air quality, motion, temperature and humidity, and light - connected via the Grove Pi hat [52]. Data was collected over a period of three months, during which a diary was kept for 14 days to document any positive control events in the home-based setting, such as coughing, sneezing, opening a window, physical activity in the room, and turning heating on/off. The collected data was uploaded to both a realtime Grafana dashboard and the RADAR-Base platform for historical analysis.

### C. Results

Figure 8 shows one of the deployments on the framework on a Raspberry Pi 4B [34] with a Grove Pi hat [52] and 3 plugged-in sensors. This is the first deployment that was installed in the shared office space.

**Fig. 8:**
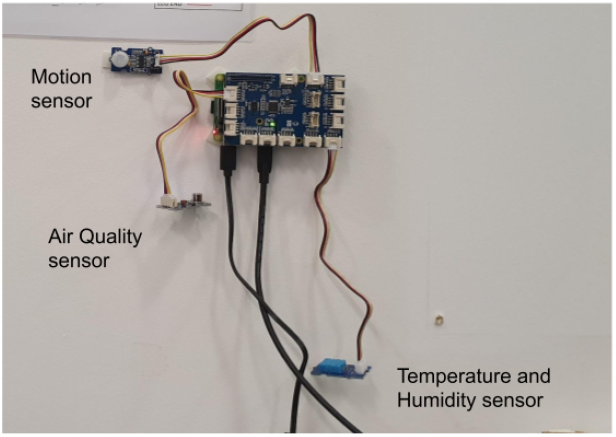
Lab deployment of the RADAR-IoT framework on a Raspberry Pi 4B [34] and Grove Pi hat [52] with 3 sensors (top: Motion Sensor, middle: Air Quality Sensor and bottom: Temperature and Humidity sensor)

Figure 9 shows the dashboard screen for a week of data collected from the above office deployment.

**Fig. 9:**
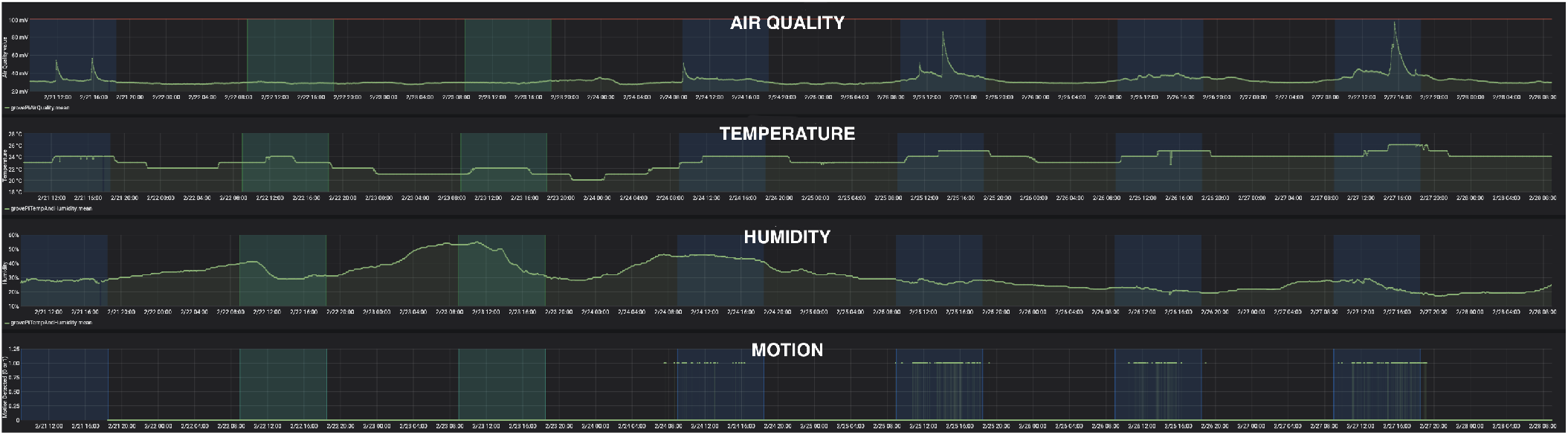
Real-time Dashboard. From top to bottom the sensors are Air Quality, Temperature, Humidity and Light. The green areas are times of day (9:00-18:00) during the weekends (holiday) while blue areas are times of day (9:00-18:00) during the weekdays. As expected the values differ between the weekdays and weekends for almost all the data due to variations in room population. For instance, there is higher Motion, Air quality and temperature during the weekdays since people are working in the room while lower humidity as compared to the weekends.

After pre-processing the data (excluding outliers) and taking the days with at least one “open-window” diary event, the correlation between the positive diary event of “open-window” with the “air-quality” sensor values was found to be on average **-0**.**61** using the spearman correlation coefficient [53]. This negative correlation is expected since opening the window will improve the air quality in the room and result in decrease in air pollutants.

The Light sensor data collected over a week of monitoring is presented in Figure 10. The plot shows expected patterns of light values, with higher values recorded in the morning when the sun rises and the room receives natural light. As the sun starts to set, there is a gradual decrease in the amount of light recorded by the sensor. When the artificial light in the room is turned on, the values jump steeply to form a second smaller peak (to the right side of the sunlight peak). The red line in the plot marks the local time of 10:30 PM, which is approximately when the user of the room goes to sleep and turns off the light, resulting in the light sensor values dropping to near **0**.

**Fig. 10:**
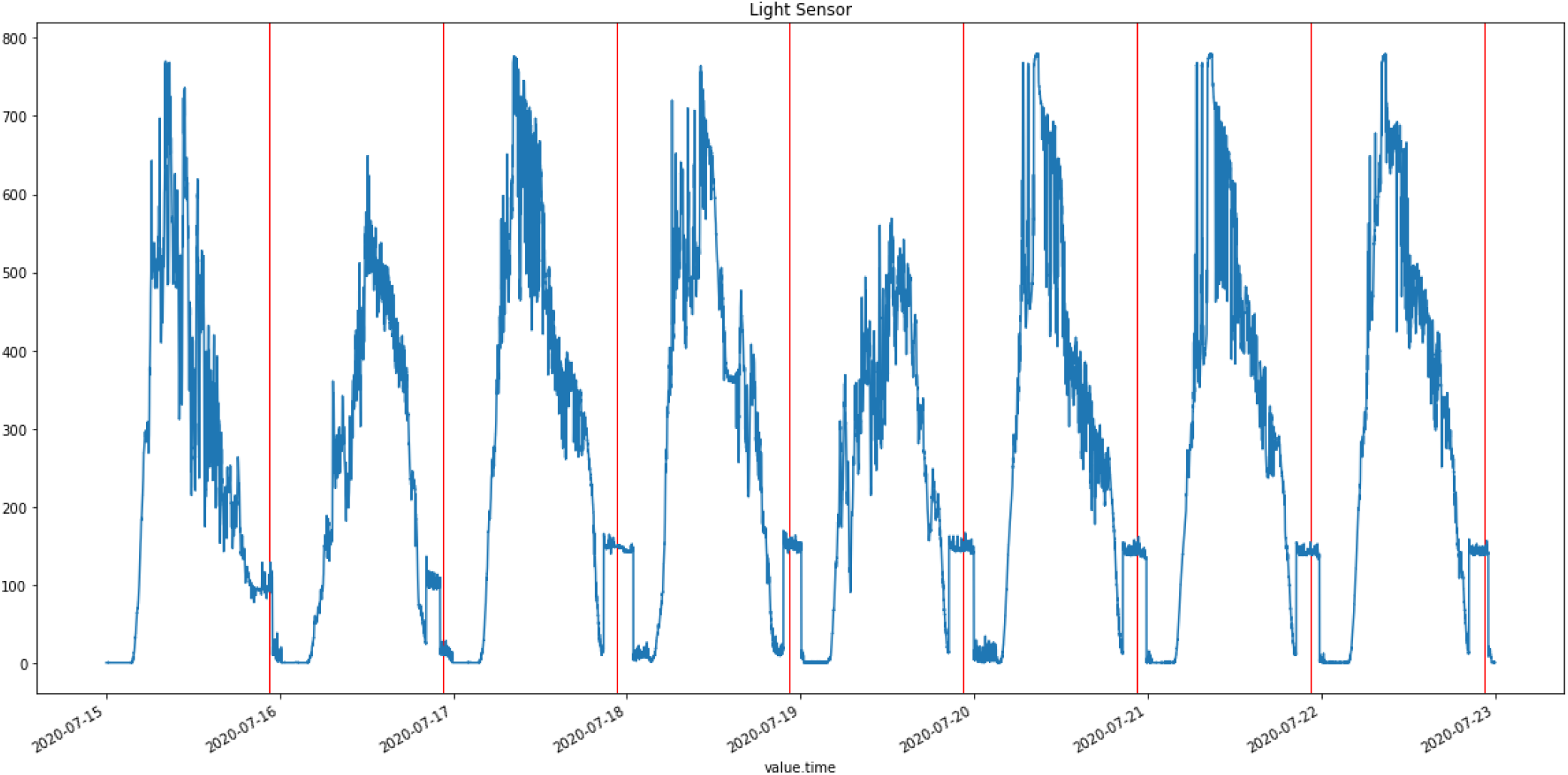
Light Sensor Data. Intraday variance during the day can be seen corresponding to sunrise and sunset.

The data collection apparatus was left on for a period of approximately 3 months to measure the completion of data. Monitoring of the setup was limited to biweekly checks, with only one issue reported during the monitoring period, which was due to a change in the home’s Wi-Fi network. The RADAR-IoT framework employed in this study has a caching mechanism that enables it to store data locally when the connection to the backend or cloud is unavailable. This allows for the caching of as much data as the available disk space permits, which can then be uploaded to the backend once the connection is restored. Table II presents the completion metrics for the sensors over the 3-month period, providing valuable insights into the reliability and performance of the data collection system.

**TABLE II:** Completion of data from home-based monitoring.

| Sensor | Percent Completion (%) | Sampling Interval (Second) |
| --- | --- | --- |
| Air Quality [54] | 90.94 | 5 |
| Temperature and Humidity [55] | 91.46 | 10 |
| Motion [56] | 83.58 | 1 |
| Light [57] | 86.59 | 30 |

All the data from the home-based setting (including the events diary data) is included here. The data is pseudonymised according to the policies and processes of the RADAR-Base platform which provides pseudonymised sensor data.

### D. Ethics

The ethical approval for this proof-of-concept was waived by the Research Ethics Committee (REC) of King’s College London (KCL) as there was no collection of primary data from human participants.

## V. Value Addition by RADAR-IoT

Using the RADAR-IoT framework with the RADAR-Base mHealth platform provides a 360° view into a user’s health and environment combining static IoT sensors dependent on an edge device and ambulatory wearable devices, providing rich information including data from:

- Wearable devices.
- Mobile phone sensors and interactions.
- Questionnaires and tasks.
- Environmental sensors like indoor air quality, temperature, humidity and toxic gas levels and many other sensor classes.

We discuss some potential application areas of monitoring using the RADAR-IoT system, this data can also be sent to the RADAR-Base mHealth platform to combine wearable data for the purposes of generating digital biomarkers.

Managing infection control can be challenging in hospital wards, monitoring systems could be used to track room visits and monitor coughing, sneezing and even particulate levels in rooms to give a readout of the status of air quality or the dynamics of airflow through a building.

Continuous passive monitoring of cough, sneezing, and snoring (including apnea and hypopnea) could be monitored using microphone and air sensors. This is interesting especially where the environment contributes directly to the disease and symptoms. For instance in the case of chronic pulmonary disorders, poor air quality can have an adverse effect on the user’s health including increased duration and intensity of coughing and wheezing. It was also shown [58] that poor air quality can even be an important factor in depression and anxiety relapse. Similar studies show that air quality improved after introduction of the London Ultra Low Emission Zones (ULEZ) [59]. These were introduced due to high air pollution in the city cause health issues for residents [60].

Another example use case is for monitoring of conditions where the participants impaired motor control or cognitive ability affect the capacity for everyday living function such as in dementias such as Alzheimer’s disease [61]. In such cases, certain environmental events can be captured using the RADAR-IoT framework which could help in understanding the progression of disease or loss of activities of daily living e.g. using door or motion sensors to detect when a participant opens a door or cupboard. Using a range of different sensing capabilities to monitor to build up a picture of home-based utility while simultaneously complementing this with wearable sensors to provide insight into their behaviour and physiology. With an increasingly elderly population, improved in-community care for patient populations will be a significant requirement, this type of continuous, passive monitoring and risk assessment or critical event detection could be incorporated into and augment a future care system.

## VI. Conclusion

RADAR-IoT takes inspiration from existing work in IoMT and seeks to address a number of limitations with current IoT gateway frameworks. These include being closed source, limited number or type of sensors and I/O protocols, restricted interoperability and scalability, no demonstrable support of cloud platforms and limited demonstration of proof-of-concept and integration with a cloud system that is well-tested in the real-world [1] [2] [17] [62].

In the design and proof of concept, deployment highlights a number of important areas where RADAR-IoT differentiates itself from related systems. Firstly, for interoperability, the system is device, sensor and programming language agnostic. The open-source nature of RADAR-IoT makes it readily extensible and the modular design allows for ease of composition. Lastly, it supports a wide range of IoT input-output protocols and multiple data sinks, including but not limited to, the RADAR-base mHealth platform, on-device AI and ML, and the Grafana dashboard. On the security and privacy side, RADAR-IoT supports industry-leading security and medical-level privacy using standards such as identity management, OAuth 2.0 [45], encryption at REST, pseudo-anonymisation and data schematisation and typing. Integration with RADAR-base [20] provides a well-established open-source mHealth cloud backend, for data collection, aggregation, transformation and more compute-intensive analytics and a combination of different types of data sources like wearables, IoT sensors, mobile apps and eCRFs.

The RADAR-IoT framework provides an edge gateway framework for integrating IoT sensors into existing cloud backends while also supporting the ability to be used on isolated local networks such as within a hospital. With the rapidly evolving digital healthcare space, there is an excellent opportunity for sensing capabilities and automation of IoT in Healthcare. This combined with the diverse challenges and the increasing gap of technologies [62] providing solutions, the RADAR-IoT framework provides a novel open-source architecture for those looking to implement IoT to research or clinical work in healthcare.

We demonstrated a proof of concept using RADAR-IoT to flexibly connect a range of IoT sensors and how this data could be collected and used with various 3rd party integrations. This showed the system and data collection operating as expected with sensors responding appropriately to changes in the environment. The RADAR-IoT framework achieved a major breakthrough by adding cutting-edge IoT data collection capabilities to the already widely-used RADAR-Base mHealth platform’s ambulatory health monitoring system. By incorporating static IoT sensors, the platform now provides a comprehensive 360-degree view of both patients’ health and environment, allowing for a more holistic understanding of how these factors interact and impact patient outcomes. This innovation has the potential to revolutionize the field of health monitoring and bring about significant improvements in patient care.

RADAR-IoT provides a reference implementation of the architecture that is a good starting point for the implementation of IoT in healthcare and beyond. These open-source data collection capabilities are a key part of the next generation of research and healthcare automation and responsive infrastructure. While the platform is novel and works well in a real-world setting, some limitations exist: these include limited sensor implementations currently, limited support for unique device types other than sensors (like audio, video), required technical knowledge and lack of a remote sensor catalog (a repository of all the sensor implementations for RADAR-IoT). Future work on the RADAR-IoT will involve mitigating these limitations. These improvements include a full gateway with both the software framework and the hardware device bundled solution for ease of use. This can also be pre-configured with sensors, e-sims, battery pack and software components based on disease types for a more plug-and-play solution. Moreover, to fully demonstrate the practical value of the framework, it is imperative to conduct further pilot studies that evaluate its performance in real-world use cases. These studies should involve detailed analysis of data collected from wearables, apps, and IoT sensors, and should aim to provide concrete evidence of the framework’s effectiveness and utility in improving patient outcomes.

## Data Availability

All data produced in the present study are available upon reasonable request to the authors

https://github.com/RADAR-base/RADAR-IoT

## VII. Acknowledgements

Backend infrastructure facilities were provided by King’s College London Open Stack Cloud “Rosalind.” Rosalind was funded with the following capital equipment grants: GSTT Charity reference: STR130505 and Maudsley Charity reference 980. The authors receive funding support from the NIHR Biomedical Research Centre at South London and Maudsley NHS Foundation Trust and King’s College London. The views expressed are those of the authors and not necessarily those of the NHS, the NIHR, or the Department of Health. The authors also acknowledge the support of NIHR University College London Hospitals Biomedical Research Centre.

## References

[1] F. Alshehri and G. Muhammad, “A comprehensive survey of the internet of things (iot) and ai-based smart healthcare,” IEEE Access, vol. 9, p. 3660–3678, 2021.

[2] S. M. R. Islam, D. Kwak, M. H. Kabir, M. Hossain, and K.-S. Kwak, “The internet of things for health care: A comprehensive survey,” IEEE Access, vol. 3, p. 678–708, 2015.

[3] “Internet of Things in Healthcare Market Size Report, 2030.” [Online]. Available: https://www.grandviewresearch.com/industry-analysis/internet-of-things-iot-healthcare-market

[4] D. Biran Achituv and L. Haiman, “Physicians’ attitudes toward the use of IoT medical devices as part of their practice,” Online Journal of Applied Knowledge Management (OJAKM), vol. 4, no. 2, p. 128–145, 2016. [Online]. Available: http://www.iiakm.org/ojakm/articles/2016/OJAKMvolume42pp128-145.php

[5] “IoT Edge Framework.” [Online]. Available: https://esf.eurotech.com/docs/edge-computing-platform

[6] “Eclipse Kura™ Documentation.” [Online]. Available: http://eclipse.github.io/kura/docs-release-5.2/

[7] Y.-H. Lee and S. Nair, “A Smart Gateway Framework for IOT Services,” in 2016 IEEE International Conference on Internet of Things (iThings) and IEEE Green Computing and Communications (GreenCom) and IEEE Cyber, Physical and Social Computing (CPSCom) and IEEE Smart Data (SmartData), Dec. 2016, p. 107–114.

[8] “ubiworx IoT Gateway Software Framework.” [Online]. Available: https://ubiworx.com/legal/2-general/34-ubiworx-iot-gateway-software-framework

[9] T. Adesina and O. Osasona, “A Novel Cognitive IoT Gateway Framework: Towards a Holistic Approach to IoT Interoperability,” May 2019.

[10] “101 Overview.” [Online]. Available: https://docs.devicehive.com/docs

[11] “ThingSpeak Documentation -MathWorks United Kingdom.” [Online]. Available: https://uk.mathworks.com/help/thingspeak/index.html?stid=CRUXlftnav

[12] “OVERVIEW.” [Online]. Available: https://docs.thinger.io/

[13] “Home . zettajs/zetta Wiki.” [Online]. Available: https://github.com/zettajs/zetta|

[14] “OpenRemote The 100% Open Source IoT Platform.” [Online]. Available: https://www.openremote.io/

[15] “ThingsBoard IoT Gateway,” Nov. 2022, original-date: 2017-01-05T05:41:03Z. [Online]. Available: https://github.com/thingsboard/thingsboard-gateway

[16] Y. Ranjan, H. Sankesara, P. Conde, J. Chang, Z. Rashid, R. J. Dobson, and A. A Folarin, “Radar-iot: An open-source, interoperable and extensible iot gateway framework for healthcare and beyond,” in Adjunct Proceedings of the 2022 ACM International Joint Conference on Pervasive and Ubiquitous Computing and the 2022 ACM International Symposium on Wearable Computers, ser. UbiComp/ISWC ‘22 Adjunct. New York, NY, USA: Association for Computing Machinery, 2023, p. 102–104. [Online]. Available: 10.1145/3544793.3560318

[17] M. A. Razzaque, M. Milojevic-Jevric, A. Palade, and S. Clarke, “Middleware for Internet of Things: A Survey,” IEEE Internet of Things Journal, vol. 3, no. 1, p. 70–95, Feb. 2016, conference Name: IEEE Internet of Things Journal.|

[18] “IoT Medical Device Gateway -Remote Patient Monitoring Lantronix,” Jul. 2014. [Online]. Available: https://www.lantronix.com/solutions/healthcare/

[19] S. Kesavan and G. K. Kalambettu, “IOT enabled comprehensive, plug and play gateway framework for smart health,” in 2018 Second International Conference on Advances in Electronics, Computers and Communications (ICAECC), Feb. 2018, p. 1–5.

[20] Y. Ranjan, Z. Rashid, C. Stewart, P. Conde, M. Begale, D. Verbeeck, S. Boettcher, T. Hyve, R. Dobson, A. Folarin, and T. R.-C. Consortium, “RADAR-Base: Open Source Mobile Health Platform for Collecting, Monitoring, and Analyzing Data Using Sensors, Wearables, and Mobile Devices,” JMIR mHealth and uHealth, vol. 7, no. 8, p. e11734, Aug. 2019, company: JMIR mHealth and uHealth Distributor: JMIR mHealth and uHealth Institution: JMIR mHealth and uHealth Label: JMIR mHealth and uHealth Publisher: JMIR Publications Inc., Toronto, Canada. [Online]. Available: https://mhealth.jmir.org/2019/8/e11734

[21] “Case Studies.” [Online]. Available: https://radar-base.org/case-studies/

[22] G. Dalla Costa, L. Leocani, X. Montalban, A. I. Guerrero, P. S. Sørensen, M. Magyari, R. J. B. Dobson, N. Cummins, V. A. Narayan, M. Hotopf, G. Comi, and on behalf of the RADAR-CNS consortium, “Real-time assessment of COVID-19 prevalence among multiple sclerosis patients: a multicenter European study,” Neurological Sciences, vol. 41, no. 7, p. 1647–1650, Jul. 2020. [Online]. Available: 10.1007/s10072-020-04519-x

[23] S. Sun, A. A. Folarin, Y. Ranjan, Z. Rashid, P. Conde, C. Stewart, N. Cummins, F. Matcham, G. D. Costa, S. Simblett, L. Leocani, F. Lamers, P. S. Sørensen, M. Buron, A. Zabalza, A. I. G. Perez, B. W. Penninx, S. Siddi, J. M. Haro, I. Myin-Germeys, A. Rintala, T. Wykes, V. A. Narayan, G. Comi, M. Hotopf, R. J. Dobson, and R.-C. Consortium, “Using Smartphones and Wearable Devices to Monitor Behavioral Changes During COVID-19,” Journal of Medical Internet Research, vol. 22, no. 9, p. e19992, Sep. 2020, company: Journal of Medical Internet Research Distributor: Journal of Medical Internet Research Institution: Journal of Medical Internet Research Label: Journal of Medical Internet Research Publisher: JMIR Publications Inc., Toronto, Canada. [Online]. Available: https://www.jmir.org/2020/9/e19992

[24] E. Bruno, A. Biondi, S. Bottcher, G. Vertes, R. Dobson, A. Folarin, Y. Ranjan, Z. Rashid, N. Manyakov, A. Rintala, I. Myin-Germeys, S. Simblett, T. Wykes, A. Stoneman, A. Little, S. Thorpe, S. Lees, A. Schulze-Bonhage, and M. Richardson, “Remote Assessment of Disease and Relapse in Epilepsy: Protocol for a Multicenter Prospective Cohort Study,” JMIR Research Protocols, vol. 9, no. 12, p. e21840, Dec. 2020, company: JMIR Research Protocols Distributor: JMIR Research Protocols Institution: JMIR Research Protocols Label: JMIR Research Protocols Publisher: JMIR Publications Inc., Toronto, Canada. [Online]. Available: https://www.researchprotocols.org/2020/12/e21840

[25] Y. Zhang, A. A. Folarin, S. Sun, N. Cummins, R. Bendayan, Y. Ranjan, Z. Rashid, P. Conde, C. Stewart, P. Laiou, F. Matcham, K. M. White, F. Lamers, S. Siddi, S. Simblett, I. Myin-Germeys, A. Rintala, T. Wykes, J. M. Haro, B. W. Penninx, V. A. Narayan, M. Hotopf, R. J. Dobson, and R.-C. Consortium, “Relationship Between Major Depression Symptom Severity and Sleep Collected Using a Wristband Wearable Device: Multicenter Longitudinal Observational Study,” JMIR mHealth and uHealth, vol. 9, no. 4, p. e24604, Apr. 2021, company: JMIR mHealth and uHealth Distributor: JMIR mHealth and uHealth Institution: JMIR mHealth and uHealth Label: JMIR mHealth and uHealth Publisher: JMIR Publications Inc., Toronto, Canada. [Online]. Available: https://mhealth.jmir.org/2021/4/e24604

[26] M. Muurling, C. de Boer, R. Kozak, D. Religa, I. Koychev, H. Verheij, V. J. M. Nies, A. Duyndam, M. Sood, H. Frohlich, K. Hannesdottir, G. Erdemli, F. Lucivero, C. Lancaster, C. Hinds, T. G. Stravopoulos, S. Nikolopoulos, I. Kompatsiaris, N. V. Manyakov, A. P. Owens, V. A. Narayan, D. Aarsland, P. J. Visser, M. Buegler, R. Fischer, R. Harms, I. B. Meier, I. Tarnanas, A. Diaz, J. Georges, D. Gove, C. de Boer, M. Muurling, P. J. Visser, I. Kompatsiaris, I. Lazarou, L. Mpaltadoros, S. Nikolopoulos, A. Papastergiou, T. Stavropoulos, D. Strantsalis, H. Froehlich, M. Hoffman-Apitius, M. Sood, N. Manyakov, V. A. Narayan, J. G. Novak, D. Religa, E. Schwertner, J. Secnik, B. Winblad, D. Aarsland, P. Conde, A. Folarin, G. Lavelle, A. P. Owens, A. McCarthy, A. Nickerson, J. Boere, B. Consiglio, Y. Daskalova, A. Duyndam, I. Kanter-Schlifke, V. J. M. Nies, P. Stolk, H. Verheij, N. Coello, J. Curcic, G. Erdemli, T. Hache, K. Hannesdottir, A. Sverdlov, V. Vallejo, E. Yang, A. Dowling, R. Kozak, M. Naylor, R. P. dos Reis, G. Shin, J. Borgdorff, E. Cirillo, K. Hedayati, N. Mahasivam, A. Doherty, C. Hinds, I. Koychev, C. Lancaster, S. Libert, F. Lucivero, Y. Wu, A. Durudas, and the RADAR-AD Consortium, “Remote monitoring technologies in Alzheimer’s disease: design of the RADAR-AD study,” Alzheimer’s Research & Therapy, vol. 13, no. 1, p. 89, Apr. 2021. [Online]. Available: 10.1186/s13195-021-00825-4

[27] Y. Zhang, A. A. Folarin, S. Sun, N. Cummins, Y. Ranjan, Z. Rashid, P. Conde, C. Stewart, P. Laiou, F. Matcham, C. Oetzmann Lamers, S. Siddi, S. Simblett, A. Rintala, D. C. Mohr Myin-Germeys, T. Wykes, J. M. Haro, B. W. J. H. Penninx, V. A. Narayan, P. Annas, M. Hotopf, R. J. B. Dobson, and R.-C. Consortium, “Predicting Depressive Symptom Severity Through Individuals’ Nearby Bluetooth Device Count Data Collected by Mobile Phones: Preliminary Longitudinal Study,” JMIR mHealth and uHealth, vol. 9, no. 7, p. e29840, Jul. 2021, company: JMIR mHealth and uHealth Distributor: JMIR mHealth and uHealth Institution: JMIR mHealth and uHealth Label: JMIR mHealth and uHealth Publisher: JMIR Publications Inc., Toronto, Canada. [Online]. Available: https://mhealth.jmir.org/2021/7/e29840

[28] S. Liu, J. Han, E. L. Puyal, S. Kontaxis, S. Sun, P. Locatelli, J. Dineley, F. B. Pokorny, G. D. Costa, L. Leocani, A. I. Guerrero, C. Nos, A. Zabalza, P. S. Sørensen, M. Buron, M. Magyari, Y. Ranjan, Z. Rashid, P. Conde, C. Stewart, A. A. Folarin, R. J. Dobson, R. Bailon, S. Vairavan, N. Cummins, V. A. Narayan, M. Hotopf, G. Comi, B. Schuller, and R.-C. Consortium, “Fitbeat: COVID-19 estimation based on wristband heart rate using a contrastive convolutional auto-encoder,” Pattern Recognition, vol. 123, p. 108403, Mar. 2022. [Online]. Available: https://www.sciencedirect.com/science/article/pii/S0031320321005793

[29] C. Stewart, Y. Ranjan, P. Conde, Z. Rashid, H. Sankesara, X. Bai, R. J. B. Dobson, and A. A. Folarin, “Investigating the Use of Digital Health Technology to Monitor COVID-19 and Its Effects: Protocol for an Observational Study (Covid Collab Study),” JMIR Research Protocols, vol. 10, no. 12, p. e32587, Dec. 2021, company: JMIR Research Protocols Distributor: JMIR Research Protocols Institution: JMIR Research Protocols Label: JMIR Research Protocols Publisher: JMIR Publications Inc., Toronto, Canada. [Online]. Available: https://www.researchprotocols.org/2021/12/e32587

[30] Y. Ranjan, M. Althobiani, J. Jacob, M. Orini, R. J. Dobson, J. Porter, J. Hurst, and A. A. Folarin, “Remote Assessment of Lung Disease and Impact on Physical and Mental Health (RALPMH): Protocol for a Prospective Observational Study,” JMIR Research Protocols, vol. 10, no. 10, p. e28873, Oct. 2021, company: JMIR Research Protocols Distributor: JMIR Research Protocols Institution: JMIR Research Protocols Label: JMIR Research Protocols Publisher: JMIR Publications Inc., Toronto, Canada. [Online]. Available: https://www.researchprotocols.org/2021/10/e28873

[31] P. Laiou, D. A. Kaliukhovich, A. A. Folarin, Y. Ranjan, Z. Rashid, P. Conde, C. Stewart, S. Sun, Y. Zhang, F. Matcham, A. Ivan, G. Lavelle, S. Siddi, F. Lamers, B. W. Penninx, J. M. Haro, P. Annas, N. Cummins, S. Vairavan, N. V. Manyakov, V. A. Narayan, R. J. Dobson, M. Hotopf, and Radar-CNS, “The Association Between Home Stay and Symptom Severity in Major Depressive Disorder: Preliminary Findings From a Multicenter Observational Study Using Geolocation Data From Smartphones,” JMIR mHealth and uHealth, vol. 10, no. 1, p. e28095, Jan. 2022, company: JMIR mHealth and uHealth Distributor: JMIR mHealth and uHealth Institution: JMIR mHealth and uHealth Label: JMIR mHealth and uHealth Publisher: JMIR Publications Inc., Toronto, Canada. [Online]. Available: https://mhealth.jmir.org/2022/1/e28095

[32] K. M. White, F. Matcham, D. Leightley, E. Carr, P. Conde, E. Dawe-Lane, Y. Ranjan, S. Simblett, C. Henderson, and M. Hotopf, “Exploring the Effects of In-App Components on Engagement With a Symptom-Tracking Platform Among Participants With Major Depressive Disorder (RADAR-Engage): Protocol for a 2-Armed Randomized Controlled Trial,” JMIR Research Protocols, vol. 10, no. 12, p. e32653, Dec. 2021, company: JMIR Research Protocols Distributor: JMIR Research Protocols Institution: JMIR Research Protocols Label: JMIR Research Protocols Publisher: JMIR Publications Inc., Toronto, Canada. [Online]. Available: https://www.researchprotocols.org/2021/12/e32653

[33] “Publish–subscribe pattern,” Feb. 2023, page Version ID: 1141729418. [Online]. Available: https://en.wikipedia.org/w/index.php?title=Publish%E2%80%93subscribe pattern&oldid=1141729418

[34] R. P. Ltd, “Buy a Raspberry Pi Zero.” [Online]. Available: https://www.raspberrypi.com/products/raspberry-pi-zero/

[35] S. Liang, C.-Y. Huang, and T. Khalafbeigi, “OGC SensorThings API Part 1: Sensing,” Jul. 2016, publisher: Open Geospatial Consortium. [Online]. Available: https://docs.opengeospatial.org/is/15-078r6/15-078r6.html

[36] “What is an IoT Gateway and How Do I Keep it Secure?” Nov. 2020. [Online]. Available: https://www.globalsign.com/en/blog/what-is-an-iot-gateway-device

[37] “InfluxDB Time Series Platform,” Jan. 2023. [Online]. Available: https://www.influxdata.com/products/influxdb/

[38] “grafana/grafana,” May 2023, original-date: 2013-12-11T15:59:56Z. [Online]. Available: https://github.com/grafana/grafana

[39] “Redis.” [Online]. Available: https://redis.io/

[40] “MQTT -The Standard for IoT Messaging.” [Online]. Available: https://mqtt.org/

[41] “Node-RED,” May 2023, original-date: 2013-09-05T13:30:47Z. [Online]. Available: https://github.com/node-red/node-red

[42] “Apache Kafka.” [Online]. Available: https://kafka.apache.org/

[43] Z. Gu, Y. Wang, W. Shi, Z. Tian, K. Ren, and F. C. M. Lau, “A Practical Neighbor Discovery Framework for Wireless Sensor Networks,” Sensors, vol. 19, no. 8, p. 1887, Jan. 2019, number: 8 Publisher: Multidisciplinary Digital Publishing Institute. [Online]. Available: https://www.mdpi.com/1424-8220/19/8/1887

[44] “Object-oriented programming,” May 2023, page Version ID: 1154120068. [Online]. Available: https://en.wikipedia.org/w/index.php? title=Object-oriented programming&oldid=1154120068

[45] “OAuth 2.0 — OAuth.” [Online]. Available: https://oauth.net/2/

[46] “Documentation.” [Online]. Available: https://avro.apache.org/docs/

[47] “Schema Registry,” May 2023, original-date: 2014-12-09T22:38:11Z. [Online]. Available:https://github.com/confluentinc/schema-registry

[48] “Nebula . Docker orchestrator for IoT & distributed systems.” [Online]. Available: https://nebula-orchestrator.github.io/

[49] “Dataplicity.” [Online]. Available: https://www.dataplicity.com/

[50] “netdata/netdata: Real-time performance monitoring, done right! https://www.netdata.cloud. [Online]. Available: https://github.com/netdata/netdata

[51] “Google Cloud IoT Core documentation Cloud IoT Core Documentation.” [Online]. Available: https://cloud.google.com/iot/docs

[52] “GrovePi Plus Seeed Studio Wiki,” Jan. 2023. [Online]. Available: https://wiki.seeedstudio.com/GrovePiPlus/

[53] C. Spearman, “The Proof and Measurement of Association between Two Things,” The American Journal of Psychology, vol. 15, no. 1, p. 72–101, 1904, publisher: University of Illinois Press. [Online]. Available: https://www.jstor.org/stable/1412159

[54] “Grove -Air Quality Sensor v1.3 Seeed Studio Wiki,” Jan. 2023. [Online]. Available: https://wiki.seeedstudio.com/Grove-AirQualitySensorv1.3/

[55] “Grove -Temperature&Humidity Sensor (DHT11) Seeed Studio Wiki,” Jan. 2023. [Online]. Available: https://wiki.seeedstudio.com/Grove-TemperatureAndHumiditySensor/

[56] “Grove -PIR Motion Sensor Seeed Studio Wiki,” Jan. 2023. [Online]. Available: https://wiki.seeedstudio.com/Grove-PIRMotionSensor/

[57] “Grove -Light Sensor Seeed Studio Wiki,” Jan. 2023. [Online]. Available: https://wiki.seeedstudio.com/Grove-LightSensor/

[58] I. Braithwaite, S. Zhang, J. B. Kirkbride, D. P. J. Osborn, and J. F. Hayes, “Air Pollution (Particulate Matter) Exposure and Associations with Depression, Anxiety, Bipolar, Psychosis and Suicide Risk: A Systematic Review and Meta-Analysis,” Environmental Health Perspectives, vol. 127, no. 12, p. 126002, publisher: Environmental Health Perspectives. [Online]. Available: 10.1289/EHP4595

[59] L. Ma, D. J. Graham, and M. E. J. Stettler, “Has the ultra low emission zone in london improved air quality?” Environmental Research Letters, vol. 16, no. 12, p. 124001, nov 2021. [Online]. Available: 10.1088/1748-9326/ac30c1

[60] “London Air Quality Network :: Welcome to the London Air Quality Network Statistics Maps.” [On-line]. Available: https://www.londonair.org.uk/london/asp/reportdetail.asp?ReportID=2019laqnr&ReportType=LAQNAnnualReport

[61] T. G. Stavropoulos, A. Papastergiou, L. Mpaltadoros, S. Nikolopoulos, and I. Kompatsiaris, “IoT Wearable Sensors and Devices in Elderly Care: A Literature Review,” Sensors, vol. 20, no. 10, p. 2826, Jan. 2020, number: 10 Publisher: Multidisciplinary Digital Publishing Institute. [Online]. Available: https://www.mdpi.com/1424-8220/20/10/2826

[62] J. Calvillo-Arbizu, I. Roman-Martinez, and J. Reina-Tosina, “Internet of things in health: Requirements, issues, and gaps,” Computer Methods and Programs in Biomedicine, vol. 208, p. 106231, Sep. 2021. [Online]. Available: https://www.sciencedirect.com/science/article/pii/S0169260721003059

